# Modelling strategies to organize healthcare workforce during pandemics: application to COVID-19

**DOI:** 10.1101/2020.03.23.20041863

**Authors:** Daniel Sánchez-Taltavull, Daniel Candinas, Édgar Roldán, Guido Beldi

**Affiliations:** Department for Visceral Surgery and Medicine, Bern University Hospital, University of Bern, Switzerland; ICTP – The Abdus Salam International Centre for Theoretical Physics, Trieste, Italy

## Abstract

Protection of healthcare workforce who are at increased risk to become infected is of paramount relevance for the care of patients in the setting of a pandemic such as coronavirus disease 2019 (COVID-19). The ideal organisational strategy to protect the workforce in a situation in which social distancing cannot be maintained remains to be determined. In this study, we have mathematically modelled strategies for the employment of hospital workforce with the goal to simulate health and productivity of the workers. The models were designed to determine if desynchronization of medical teams by dichotomizing the workers may protect the workforce. Our studies model workforce productivity depending on the infection rate, the presence of reinfection and the efficiency of home office and apply our theory to the case of COVID-19. The results reveal that a desynchronization strategy in which two medical teams work alternating for 7 days increases the available workforce.

## 1. Introduction

During the spread of worldwide pandemics, protecting and supporting caregivers to maintain the workforce in hospitals is a crucial and challenging task. Under such extraordinary situations, the efficiency of healthcare workforce is threatened by several factors including: 1) infected patients, 2) infected co-workers and 3) infected persons outside the hospital. This is of high importance in the current pandemic of coronavirus disease 2019 (COVID-19) given that the virus may be transmitted by asymptomatic persons based on the long incubation period averaging at 5 days (1, 2, 3). Unlike for most other professions, social distancing is typically not possible in medical teams where healthcare workers are required to work in close contact with patients and coworkers.

Currently, little is known about which work organizational strategies in the hospital are most suitable protect the healthcare workforce. Potential strategies may include prolonging duty hours and thereby limiting interactions between coworkers or complete desynchronization of the workforce in which teams are dichotomized and each half of the team works for one week alternatively.

In this short report, we perform simulations of biophysical models of coronavirus epidemics of COVID-19 in a healthcare working team, and discuss the efficiency of different work strategies during the viral outbreak.

Different types of mathematical models have been used to study epidemiology. In the classical Susceptible Infectious Recovered (SIR) model, a susceptible patient (S) can be infected (I), and the infected person can recover (R), without the risk of reinfection. Variants of this model, are the Susceptible Infectious Susceptible (SIS) and Susceptible Exposed Infectious Recovered (SEIR) models (4). The SEIR model in which the recovered patients are susceptible again, has already been used for modeling purposes during the COVID-19 outbreak (5). Also SEIR models have been used to model control of expansion in the context of the COVID-19 and have been extended to include age and asymptomatic cases (6, 7). Furthermore, SEIR models have applied in this context by including persons in quarantine (QSEIR model) (8, 9). A model based on Microscopic Markov Chain approach have been proposed to study the effect of confinement (10).

Here, we put forward SIS and SIR models by dividing the infectious persons into a latent and infected state in order to account for the potentially long asymptomatic phase of COVID-19. Next, have developed two time-dependent compartmental models with and without reinfection by adapting the SAIR model (11-13). Next, we add a variable to account for work *W* and build two mathematical models for COVID-19 described by ordinary differential equations (ODE). These are denoted as Susceptible Latent Infected Work (SLIW) model and Susceptible Latent Infected Recovered Work (SLIRW) model. In our SLIW model, the workers can be healthy and susceptible to be infected, *S*, infected in the incubation period, *L*, infected presenting symptoms, *I*. Additionally in the SLIRW model, after recovery the patients become immune to new infections, *R*. Additionally, the models are adapted to account for work done, *W*, by the non-infected workers.

To investigate the possible workforce organizations, we use ODE describing the dynamics of the models including time-dependent parameters in which the rate changes based on their location, that is, in-hospital compared with home office. Deterministic models may not be accurate to describe the dynamics when small populations are present (14). In the context of COVID-19, stochastic epidemiology models have been used to study confinement strategies (15)

Therefore, to investigate the effect of extra measures, such as extra group divisions with space separation, and testing, we reformulated our model in terms of a master equation (16).

The rest of the report is organized as follows: We study and compare organizational strategies on the hospital workforce in the situation of a pandemic on productivity in the absence of reinfection and with reinfection (section 2), and present the results of simulations (section 3). Finally, our results are summarized and discussed (section 4).

## 2. Models

### 2.1. Sketch of the models

Figure 1 shows the SLIW and SLIWR models with compartmentalization that we describe in detail in Section 2.1.2 and Section 2.2.2, respectively. In our models, we consider that a group of healthcare workers are divided in two teams of equal size. At any time, one team is at the workplace whereas the other team stays at home. A dichotomous work switch is implemented as follows (Figure 1A): during the first week Team 1 stays at the workplace and Team 2 at home; during the second week Team 1 stays at home and Team 2 at the workplace; next the teams continue alternating their location at the end of each week. When at the workplace, each healthy susceptible worker can become latent at a time-dependent infection rate that is larger than when staying at home. Latent workers develop the infection at the workplace, at the same rate than at home. Infection of a susceptible by a latent worker can only occur in the workplace. We also assume that infected workers recover at the same rate at the workplace than at home, becoming again susceptible (SLIW) or fully recovered and immune (SLIRW).

**Figure 1.**
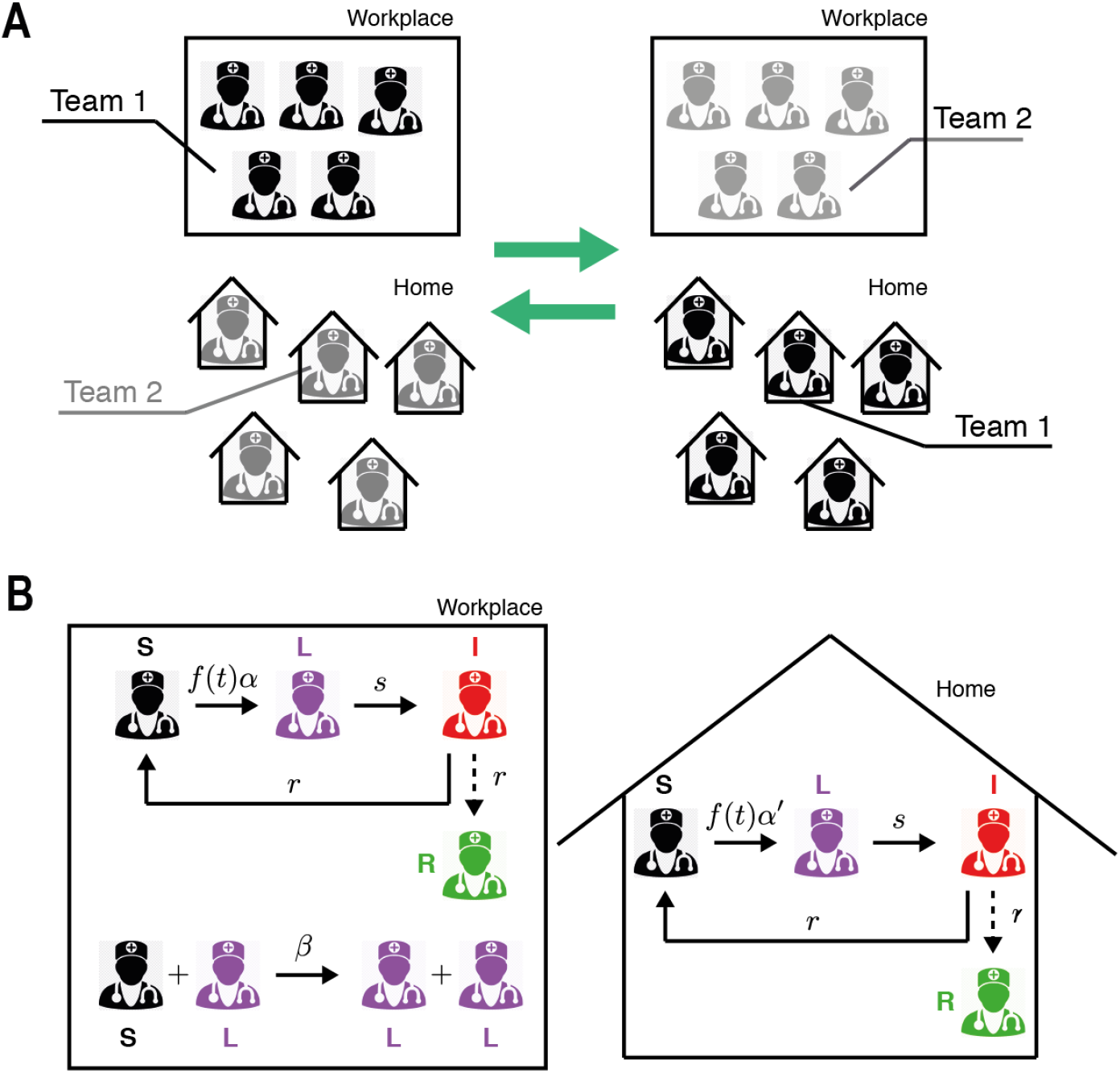
Sketch of the Susceptible Latent Infected Work model (SLIW) and Susceptible Latent Infected Recovered Work model (SLIRW) models. (A) Scheme of work shifts between two teams changing their location in the workplace and at home once per week. (B) Infection dynamics at the workplace and home. See Sec. 2.2 and 2.3 for further details. Image of the doctor adapted from (17).

### 2.2. Model with reinfection (SILW)

#### 2.2.1. One group of healthcare workers

Our first infection model describes the evolution in time *t* of the number *S* of healthy workers susceptible to be infected, the number, *L*, of infected persons in a latent state, and the number *I* of infected persons presenting symptoms. To study the impact on the workers’ efficiency, we introduce a function *W* that accounts for the amount of work done by all the available workers. The dynamics of each population are described by the following system of ODE.

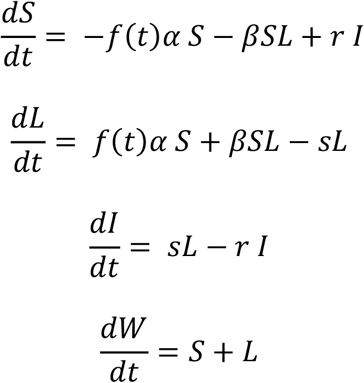

where *α* is the infection rate, resulting into an infected in a latent state. *f*(*t)* is a function accounting for the number of people infected outside the hospital, which we assume is growing in time. *β* is the infection rate when the person is infected by a co-worker, *s* is the inverse of the incubation period, or the activation rate, in which a patient in a latent state starts presenting symptoms. *r* is the recovery rate, where an infected patient recovers and becomes healthy. *W* is a function that accounts for the work output of the workers, for simplicity we assume its growth is proportional to the number of available workers. For simplicity, we consider the death rate negligible and we do not consider recoveries coming from latent patients. In our simulations reported below, we set the initial condition to *S(0)=300, L(0)=0, I(0)=0*, and *W(0)=0*, thereby representing a large unit with 300 workers.

#### 2.2.2 Two groups of healthcare workers

Next, we studied a desynchronization strategy, in which the workers are divided in half, one group works for one week in the hospital while the other works at home, and the week after they change their functions. In doing so, we model it as two groups of healthy persons susceptible to be infected, *S*_1_ and *S*_2_, two groups of infected persons in a latent state, *L*_1_ and *L*_2_, and two groups of infected persons presenting symptoms, *I*_1_ and *I*_2_. The dynamics of each population are described by the following system of ODE

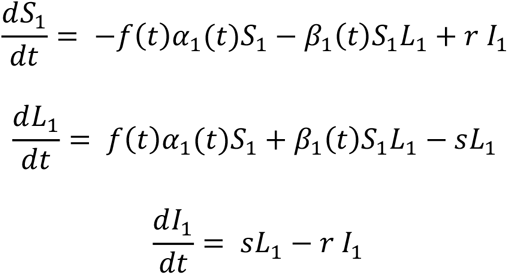

for the first group and

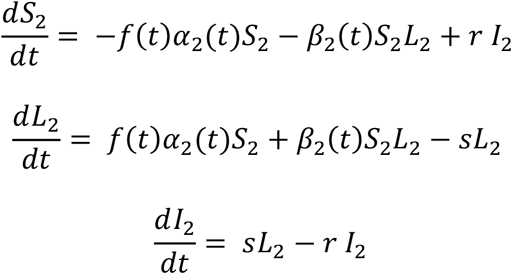

for the second group. The rate of change of the work output obeys

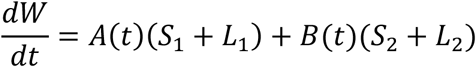

For this model, we use the same parameters than in the previous model, except for *α*_*j*_ and *β*_*j*_ (with j=1,2). Therefore we simulate our model for the first seven days with *β*_2_ = 0, *α*_2_ = 0.1*α*_1_, because the two groups are not in contact with the co-workers, and we assume that the probability to become infected is higher when working in the hospital. For the following seven days, we switched the group’s location, by exchanging their parameter values. This procedure is repeated every 7 days. We assume that the productivity decreases during home office, therefore we choose *A(t)=1* and *B(t)<A(t)* during the odd weeks and *B(t)=1* and *A(t)<B(t)* during the even weeks.

Based on the available data of COVID-19, we choose 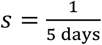, because the average duration of the incubation period is 5 days, and 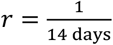 because the average recovery time is 14 days. Additionally, we chose a logistic function 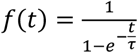 with characteristic time *τ* = 5.

### 2.3. Model without reinfection allowing for recovery (SLIRW)

#### 2.3.1. One group

In this section we study the effect of a model in which the patients develop immunity to the disease after the infection. Therefore, we add a variable for the recovered patients, R, which cannot become infected after reaching that state, and the dynamics are described by the following system of ODE

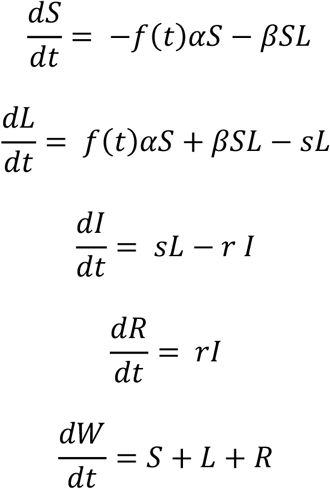

With *S(0)=300, L(0)=0, I(0)=0, R(0)=0* and *W(0)=0* as initial condition.

#### 2.3.2. Two groups

Here the desynchronization strategy is modelled for a situation in which the workers develop immunity and cannot be reinfected. This includes two groups of healthy persons susceptible to be infected, *S*_1_ and *S*_2_, two groups of infected persons in a latent state, *L*_1_ and *L*_2_, two groups of infected persons presenting symptoms, *I*_1_ and *I*_2_, and two groups of recovered persons who cannot become infected, *R*_1_ and *R*_2_,. The dynamics of each population are described by the following system of ODE, for the first group

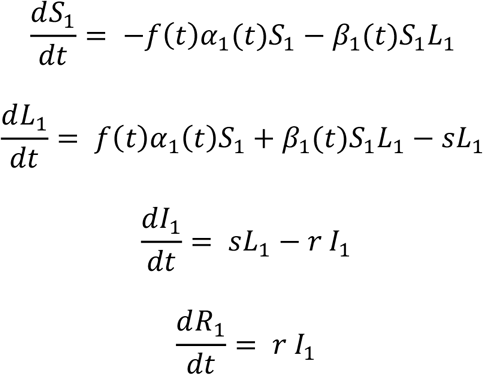

whereas for the second group

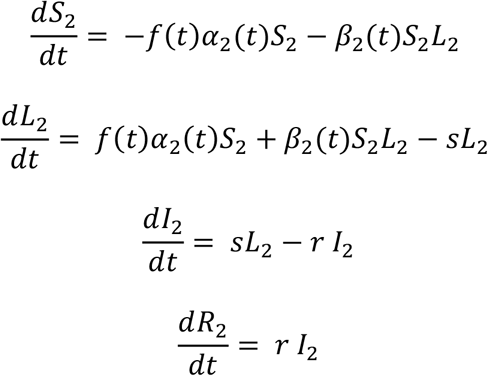

The total work done by the two groups is given by

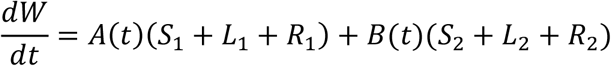

### 2.4. Stochastic models

The SLIW and SLIRW models are reformulated in terms of the following Master Equation

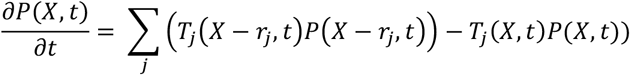

Where *P*(*X, t)* is the probability of being at state X at time t, where *X* = (*S, L, I)* in the SLIW model, *X* = (*S*_1_, *L*_1_, *I*_1_, *S*_2_, *L*_2_, *I*_2_) in the desynchronized SLIW model, *X* = (*S, L, I, R)* in the SLIRW model and *X* = (*S*_1_, *L*_1_, *I*_1_, *R*_1_, *S*_2_, *L*_2_, *I*_2_, *R*_2_) in the desynchronized SLIRW model. The transition rates, *T*_*j*_, and the state changes, *r*_*j*_, are described in Tables 1 and 2.

**Table 1.** Transition rates and state changes describing the SLIW model.

| Transition rate | State change | Description |
| --- | --- | --- |
| $T_1 = \alpha S_i$ | $r_1 = (-1, 1, 0)$ | $S_i \rightarrow L_i$ |
| $T_2 = \beta S_i L_i$ | $r_2 = (-1, 1, 0)$ | $S_i + L_i \rightarrow L_i + L_i$ |
| $T_3 = s L_i$ | $r_3 = (0, -1, 1)$ | $L_i \rightarrow I_i$ |
| $T_4 = r I_i$ | $r_4 = (1, 0, -1)$ | $I_i \rightarrow S_i$ |

**Table 2.** Transition rates and state changes describing the SLIWR model.

| Transition rate | State change | Description |
| --- | --- | --- |
| $T_1 = \alpha S_i$ | $r_1 = (-1, 1, 0, 0)$ | $S_i \rightarrow L_i$ |
| $T_2 = \beta S_i L_i$ | $r_2 = (-1, 1, 0, 0)$ | $S_i + L_i \rightarrow L_i + L_i$ |
| $T_3 = s L_i$ | $r_3 = (0, -1, 1, 0)$ | $L_i \rightarrow I_i$ |
| $T_4 = r I_i$ | $r_4 = (0, 0, -1, 1)$ | $I_i \rightarrow R_i$ |

The Work done, *W*, is described as the ODE of the deterministic models. For simplicity in the master equation formulation, we replace *f* (*t)* with 1. Our transition rates change from time to time, which could be problematic with our Gillespie simulations. To artificially reduce the step size in the Gillespie algorithm, we add a reaction ∅ → ∅ with a rate 0.01 days^-1^.

### 2.5. Spatial desynchronization

In order to model space division, we divide our variables *S*_*i*_, *L*_*i*_, *I*_*i*_, *R*_*i*_, in groups, *S*_*i,j*_, *L*_*i,j*_, *I*_*i,j*_, *R*_*i,j*_ with *j* = 1, …, *N* where *N* is the number of groups. Since the space is divided by *N* where *N* is the number of groups, we assume the rate *β* is multiplied by *N*, that is, *β* → *NB*.

## 3. Results

### 3.1. Simulations of the SLIW model

First, we study the effect of the infection rates on the health of the workers. In figure 2 we show the simulations of our systems of ODEs with two representative sets of parameters, low infection rate (Figure 2Ai) and high infection rate (Figure 2Aii). Regardless the values, we observe that the strategy of dividing the workers in two groups decrease the number of infected workers, thus increasing the available workforce. In Figure 2B we show the number of workers as a function of the infection rates. We observe that with the desynchronization strategy the number of healthy workers is higher than with the normal strategy.

**Figure 2.**
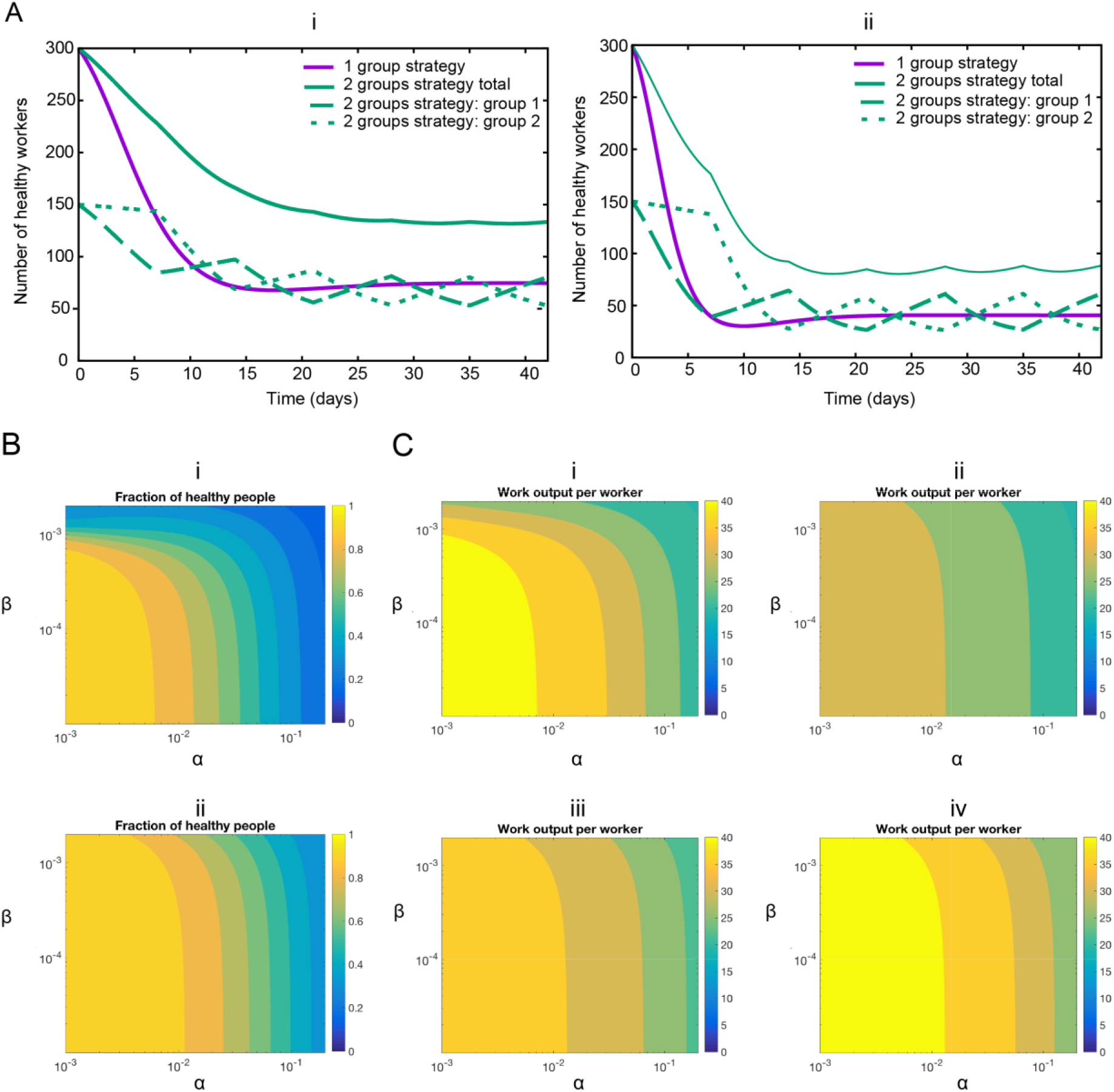
(A) Representative examples of infection rates of healthcare workers with and without desynchronization (SLIW). Number of healthy workers as a function of time for two sets of parameters values. (i) Assumption: low infection rate: *α* = 0.1, *β* = 0.001. *α*_1_ = 0.1, *β*_1_ = 0.001 during the odd weeks, and *α*_2_ = 0.1, *β*_2_ = 0.001 during the even weeks. (ii) Assumption: high infection rate: *α* = 0.2, *β* = 0.002. *α*_1_ = 0.2, *β*_1_ = 0.002 during the odd weeks, and *α*_2_ = 0.2, *β*_2_ = 0.002 during the even weeks. All rates are in units of day^-1^. (B) Impact of infection rates on healthcare workers availability. Number of healthy workers as a function of the infection rates. (i) Normal strategy. (ii) Desynchronization strategy *α* stands for *α*_1_ during the odd weeks and *α*_2_ during the even weeks, idem for *β*. The rates *α* and *β* in units of day^-1^. (C) Impact of infection rates on the productivity. Value of the work output W(t) divided by the total number of workers at the end of the simulation (6 weeks) for the normal strategy (Strategy 1), and different values of the home office productivity with the desynchronized strategy (Strategy 2). (i) Strategy 1, (ii) Strategy 2, with productivity at home B=0.5, (iii) Strategy 2, productivity at home B=0.75, (iv) Strategy 2, productivity at home B=1. The rates *α* and *β* in units of day^-1^.

So far, we have observed that the desynchronization strategy increases the number of healthy workers. Because a desynchronization strategy includes decreased productivity of home office, we next determined the overall economic impact of this strategy. Therefore, we simulate our model for 6 weeks and we represent the value of *W* divided by the total number of workers as a function *A(t)* and *B(t)* during the home office week (Figure 2C). We observe that if the productivity decreases by 50% during the home office week, the normal strategy performs better than the desynchronization strategy. For higher values (75% or 100%) of the home office productivity, the desynchronization strategy outperforms the normal strategy for high infection rates.

### 3.2. Simulations of the SLIRW model

Next, we study the effect of the infection rates in a model allowing for reinfection. In Figure 3A we show the simulations of our systems of ODE with two representative sets of parameters. Regardless the values, we observe that the strategy of dividing the workers in two groups increase the number of healthy workers during the peak of infection. In Figure 3B we show the number of minimum healthy workers as a function of the infection rates during a 6-week simulation.

**Figure 3.**
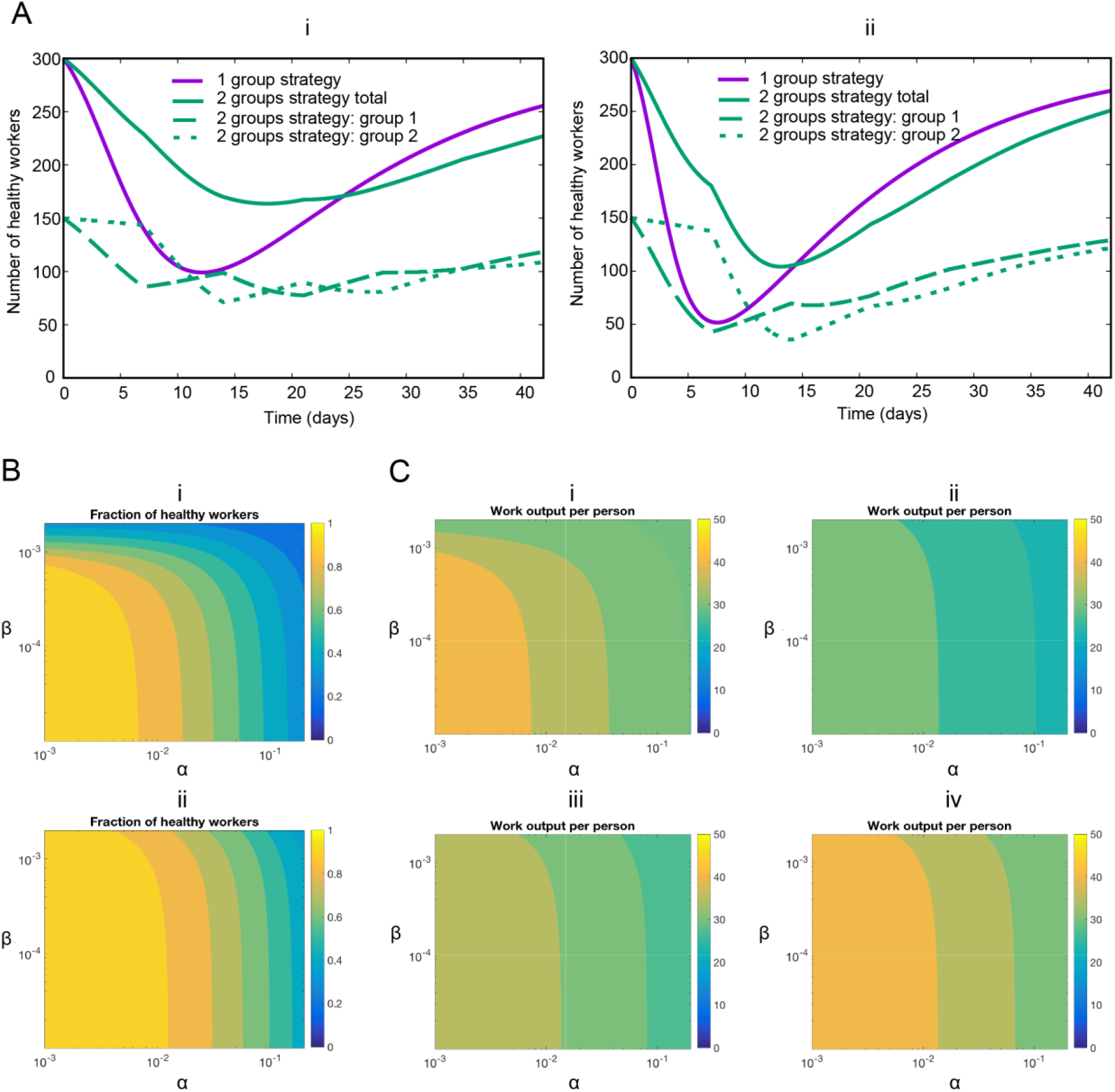
Representative examples of infection rates of healthcare workers with and without desynchronization (SLIRW). Number of healthy workers (susceptible plus recovered) as a function of time. (i) Assumption: low infection rate: *α* = 0.1, *β* = 0.001. *α*_1_ = 0.1, *β*_1_ = 0.001 during the odd weeks, and *α*_2_ = 0.1, *β*_2_ = 0.001 during the even weeks. (ii) Assumption: high infection rate: *α* = 0.2, *β* = 0.002. *α*_1_ = 0.2, *β*_1_ = 0.002 during the odd weeks, and *α*_2_ = 0.2, *β*_2_ = 0.002 during the even weeks. All rates are in units of day^-1^. (B). Impact of infection rates on healthcare workers availability during the peak infection rate. Minimum fraction of healthy workers during a simulation as a function of the infection rates. (i) Normal strategy. (ii) Desynchronization strategy *α* stands for *α*_1_ during the odd weeks and *α*_2_ during the even weeks, idem for *β*. The rates *α* and *β* in units of day^-1^. (C). Impact of infection rates on the productivity. Value of W(t) at the end of the simulation for the normal strategy (Strategy 1), and different values of the home office productivity with the desynchronized strategy. (i) Strategy 1, (ii) Strategy 2, productivity at home 0.5, (iii) Strategy 2, productivity at home 0.75, (iv) Strategy 2, productivity at home 1. The rates *α* and *β* in units of day^-1^

So far we have observed that the desynchronization strategy increases the number of healthy workers also in the situation without reinfection. To determine the economic impact, we simulate our model for 6 weeks and we represent the value of *W* as a function *α* and *β*, for different values of the home office efficiency (Figure 3C).

### 3.3. Comparison: SLIW vs SLIRW models

Next, we compared the SLIW and SLIRW. In doing so, we defined *i*_*R*_ as the infection rate and we computed the value of *W* as a function the home office efficiency for different values of the infection rate, by choosing *α* = 0.1*i*_*R*_ and *β* = 0.001*i*_*R*_ (Figure 4). We observe that for the SLIW model requires a smaller home office efficiency than for the SLIRW model.

**Figure 4.**
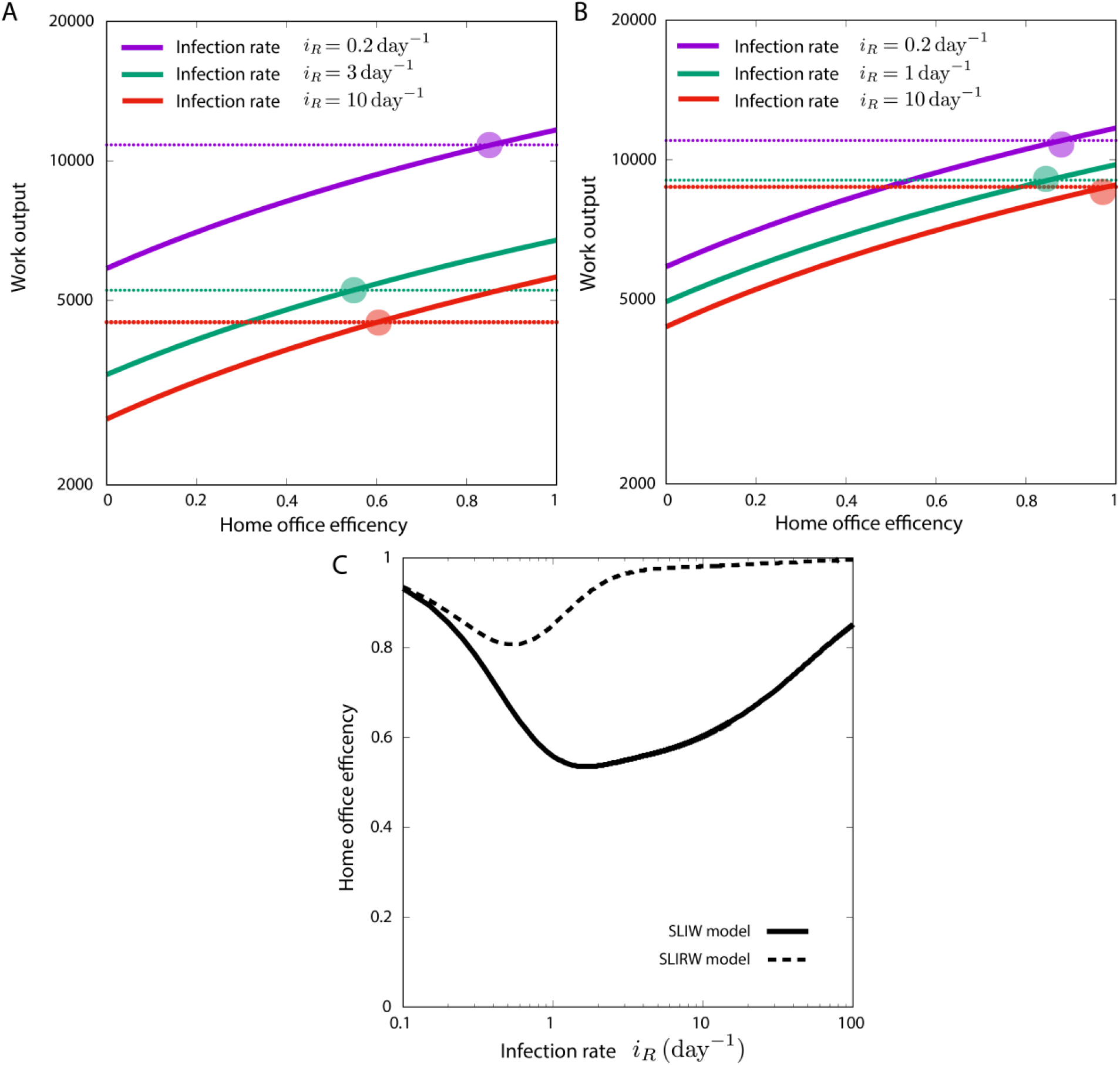
Value of W (work done) at the end of the simulation as a function of the home office efficiency for different infection rates, where *α* = 0.1*i*_*R*_ and *β* = 0.001*i*_*R*_. The dashed threshold is the productivity with the 1 group strategy. (A) SLIW model. (B) SLIRW model. (C) Value of home office efficiency *B* needed to be as productive as the normal strategy, as a function of infection rates, where *α* = 0.1*i*_*R*_ and *β* = 0.001*i*_*R*_ for SLIW and SLIWR models.

### 3.4. Spatial desynchronization

In this section we explore the effect of dividing the work space by reducing the number of persons working in a team. The workers are assigned to one small team and are never in contact with workers from other teams. In doing so, we simulate the stochastic model described in sections 2.4 and 2.5 using Gillespie simulations.

In Figure 5A,B we observe that several trajectories of the stochastic are not infected and remain at 1 and the fraction. The number of Gillespie trajectories staying at 1 increases with the number of groups, while the mean field dynamics remain the same. In Figure 5C we observe that, on average, an increase in the number of groups is translated into a higher number of healthy workers. Note that the changes in the number of groups come with an increase in *β*_*i*_ that makes the ODE system give the same result (Figure 5A). Therefore, the spatial division incorporates to our system a purely stochastic protective mechanism.

**Figure 5.**
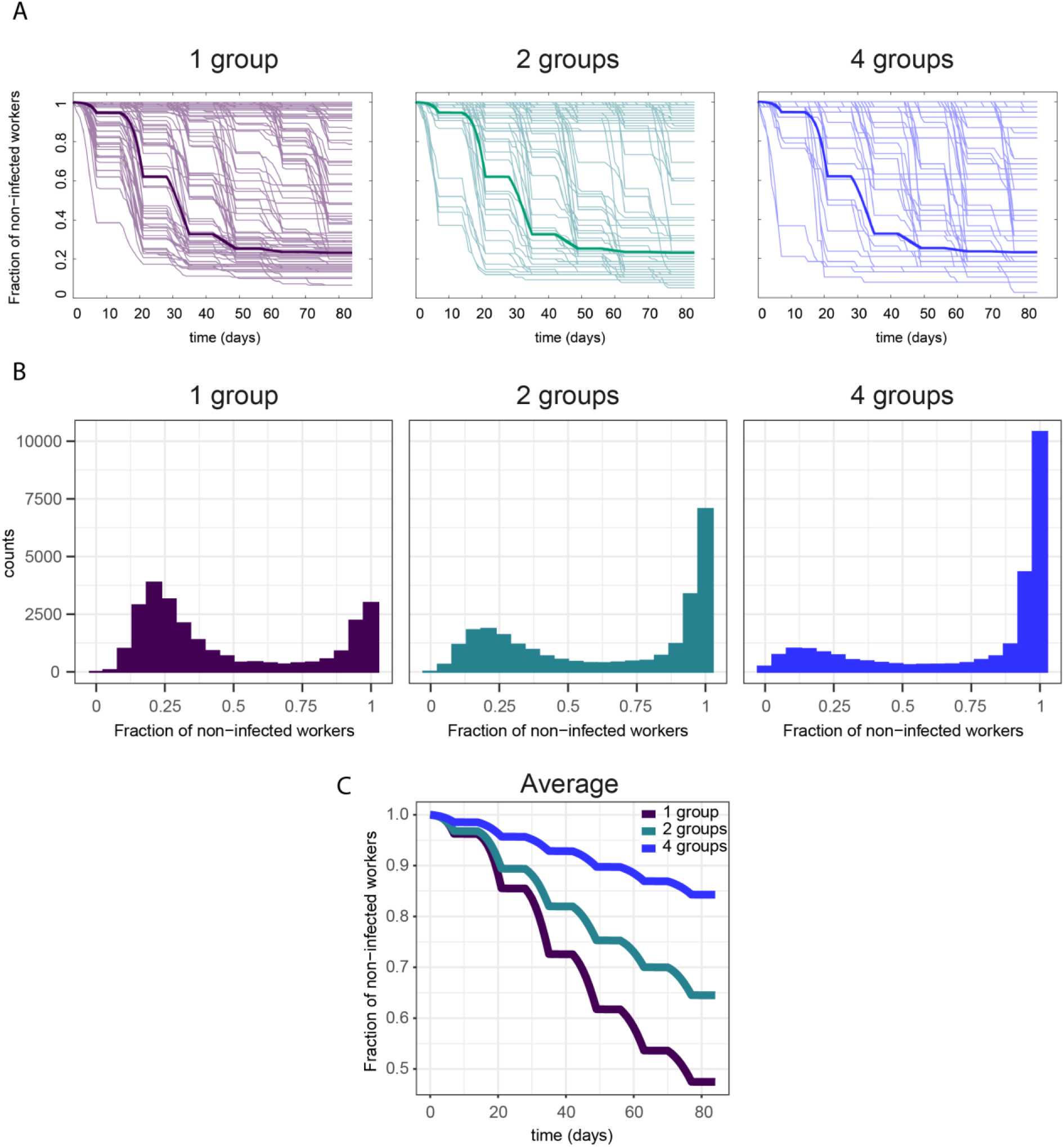
(A) Mean-field dynamics (thick line) and 100 gillespie simulations (thin lines) of *S*_*i*,1_(*t)* during a 12-week simulation of the stochastic model with spatial separation, for a group of 150 workers (1 group), a group of 75 workers (2 groups), and a group of 38 workers (4 groups). (B) Distribution of *S*_*i*,1_(*t)* at the end of a 12-week simulation, computed using 25000 gillespie simulations of the stochastic model with spatial separation, for a group of 150 workers (1 group), a group of 75 workers (2 groups) and a group of 38 workers (4 groups). (C) Average of *S*_*i*,1_(*t)* for a group of 150 workers (1 group), a group of 75 workers (2 groups) and a group of 38 workers (4 groups). (C) Average of *S*_*i*,1_(*t)* in the model. Parameters: *α*_*i*_ = 0.0005, *β*_*i*_ = 0.005 (1 group), *β*_*i*_ = 0.01 (2 groups), *β*_*i*_ = 0.01974 (4 groups).

## 4. Discussion

Given the urgent need to protect caretakers, we present here modelling approaches that address the impact of desynchronization of healthcare workforces in a short report. We have developed two time-dependent compartmental models (SLIW/SLIRW with and without reinfection) by adapting the SAIR model of (11-13). A time-dependent compartmental model was included to account for the desynchronization of healthcare teams. In addition to the availability of healthcare workers, we modelled productivity by incorporating different levels of work performed at home office.

The SLIW model showed that the desynchronization strategy is associated with an increase of the number of healthy workers compared to no desynchronization. This effect is present with both high and low levels of infection rates (Figure 2).

Next, we incorporated productivity of the workers in the period of home office. In practice, productivity of home office may depend on the tasks, which may be done outside the hospital such as writing reports. We have modelled the productivity rate of home office as a function of the infection rates. Figure 2 and figure 4 show that, for our case study model of COVID-19, a decrease of productivity to 50% during home office (only half of the time can be used for productive work) does not imply a substantial decrease of the overall productivity. However, if the productivity at home office is above 75%, overall productivity is increased with a desynchronization strategy for high infection rates.

When the SLIRW model was used to incorporate full recovery without the potential of reinfection, the protective effect of desynchronization weans over time (Figure 3). In this situation, however, the number of healthy workers with desynchronization increases, especially during the peak of infected workers (Figure 3). This observation is potentially the consequence of workers being immune to the infection thereby not requiring a desynchronization anymore. The protection of healthcare workforce (Figure 3B) and productivity (Figure 3C) also depends to the infection rate in SLIRW model.

Next, we aimed to determine the ideal level of productivity for each model (Figure 4). The ideal home office productivity strongly depends on infection rates in the model with reinfection (SLIW). In this model productivity is required to be around 60% for infection rates 3 and 10. In the model without reinfection (SLIRW), which is rather the case with COVID-19, the home office productivity needs to be higher in order to keep overall productivity similar between the one and two group strategy.

However, the way the model was designed, we artificially included a handicap to the desynchronization strategy. As long as it remains unclear if reinfection is possible, the desynchronization strategy would be maintained. Such strategy would be stopped for recovered workers if reinfection does not occur because of immunity.

We studied the stochastic effects on our model by reformulating the systems in terms of a Master Equation. Intrinsic noise is of special interest when the number of individuals is small, which was of crucial importance in our next *in silico* experiments, where we divided our workers in multiple independent groups.

The stochastic model was used to study spatial separation and revealed that for low levels of *α*_*i*_, the spatial separation reduces the spread of the disease among the workers. We investigated the origin of this effect by studying the distribution of the number of infected workers, and we have shown that most of the simulations had no infections (Figure 5A,B), reducing considerably the average number of infected workers (Figure 5C). This stochastic protective measure is a discrete effect similar to ones previously reported in biology (18)

Our model has limitations that can be addressed in multiple ways. Another extension would be to add asymptomatic patients to the model, that is, patients that pass the infection without presenting symptoms. Furthermore, the model does neither address at what costs such a desynchronization strategy would come. Given that a complete set of workers is always present, there might be losses of acquired experience for the group absent. Furthermore, these models neither address the impact of length of duty shifts nor the impact of potentially impaired communication as a consequence of decreased interactions between healthcare workers.

In summary, our model is a starting point to study how to protect healthcare workers while determining economic impact during a pandemic outbreak.

## Data Availability

All data are available in the manuscript.

## Aknowledgements

We would like to thank the Swiss National Science foundation for funding (grant number 196059).

